# COVID-19 vaccination status is associated with physical activity in German-speaking countries: the COR-PHYS-Q cohort study

**DOI:** 10.1101/2022.01.01.21268576

**Authors:** F. Schwendinger, H. T. Boeck, D. Infanger, M. Faulhaber, U. Tegtbur, A. Schmidt-Trucksäss

## Abstract

**Background:** To examine the association between COVID-19 vaccination status and physical activity (PA), sporting behavior, as well as barriers to PA in adults in Switzerland, Germany, and Austria.

**Methods:** A total of 1516 adults provided complete responses to our online questionnaire sent out in August 2021. Information about self-reported PA categories, sporting behavior, barriers to PA, and COVID-19 vaccination status were gathered. Main analyses were done using multiple linear regression adjusted for relevant parameters.

**Results:** We found a significant association of vaccination status with total PA (*p* = .011), vigorous PA (*p* = .015), and moderate PA (*p* = .001) but not transport-related PA or sedentary time. Unvaccinated adults tended to have more total and vigorous PA than those vaccinated once (ratios of geometric means: 1.34 and 1.60, respectively) or twice (1.22 and 1.09, respectively). Yet, not sufficient evidence was available to confirm this. There was no between-group difference in the contribution of leisure time, work-related, or transport-related PA to total PA. Vaccination status was not associated with sporting behavior except for jogging as the primary intensive type of sports. Finally, there were no significant differences in any of the COVID-19 specific barriers to PA between groups.

**Conclusion:** Our data showed that vaccination status is associated with PA even in summer, where the number of COVID-19 cases was low and the severity of safety measures was mild. These findings may enhance future research and improve/extend COVID-19-specific PA guidelines.

## 1. Introduction

Safety measures enacted by the governments to prevent the spread of SARS-nCoV-2 have led to impaired levels of physical activity (PA) and more sedentary time in the general population.^1^ Changes in PA patterns are an important research area as both the amount and intensity of PA are closely connected to the risk for cardiovascular disease and all-cause mortality.^2, 3^ Moreover, physical inactivity causes an enormous financial burden for healthcare systems^4^ that may have even increased during this pandemic.

There are clear recommendations by health organizations (e.g. World Health Organization^5^, US Centers for Disease Control and Prevention^6^) to carry out regular PA while abiding by public health safety measures in pandemic times. PA is a potent stimulus improving immune function, physical and mental health, as well as wellbeing.^7^ A study of 48,440 adult patients found that following PA guidelines was strongly linked to a lower risk of severe COVID-19 course.^8^ Further, Campbell & Turner ^9^ concluded that evidence from epidemiological studies points towards a physically active lifestyle lowering the incidence of communicable (e.g. bacterial and viral infections) and non-communicable diseases (e.g. cancer). This means that regular exercise improves immune competence.^9^

To date, several potent vaccines to protect against COVID-19 are available.^10-12^ From mid-January of this year (2021), mass vaccination was initiated in Switzerland, Germany, and Austria, as a result of which more than 51% of the population were fully vaccinated in August 2021.^13^ Progress with vaccinations has also led to the implementation of new regulations. Certain facilities could only be visited if individuals provided proof of either a current negative COVID-19 test result or of being fully vaccinated or recovered. These measures vary in their severity according to the number of new SARS-nCoV-2 infections and hospitalizations.

It may be speculated that fully vaccinated people and those that recovered from COVID-19 are less restricted in their PA than those who are not vaccinated. The availability of a vaccine might therefore help increase PA in the general population.^14^ Nonetheless, differences in PA might not be apparent in the summer months since the safety measures in place were looser^15^ and the number of infections was clearly below the current ones (08.08.2021, 13 cases per million people vs. 29.11.2021, 935 cases per million people^16^).

There are no studies available to date shedding light on these issues. Thus, we conducted the first transnational prospective observational study to examine the association between COVID-19 vaccination status and PA patterns, sporting behavior, and PA barriers in adults in German-speaking countries.

## 2. Methods

### 2.1 Study design

COR-PHYS-Q is a prospective observational study that is conducted at three points in time, i.e. during two weeks in August 2021, December 2021, and August 2022, respectively. The abbreviation stands for linking CORonavirus disease 2019 and PHYSical activity using a Questionnaire.

### 2.2 Setting

Data were collected using an online questionnaire distributed via e-mail, social media and popular mobile apps, websites, online-newspaper articles, and mouth-to-mouth. The present study focused on the data obtained during survey round one from 9^th^ to 23^rd^ August 2021. The project was approved by the responsible ethics committees in Switzerland (EKNZ 2021-00775), Germany (9863_BO_K_2021), and Austria (Certificate of good standing 51/2021). All procedures followed the Declaration of Helsinki.

### 2.3 Study cohort

German-speaking adults (≥18 years) living in Switzerland, Germany, or Austria were eligible for inclusion. Exclusion criteria were being unable to follow the study procedures (e.g. insufficient knowledge of the study language), mental incapacity, and individuals not having their primary residence within one of the mentioned countries. We aimed to recruit as many participants as possible and expected 2000 individuals in the three countries to fill out the questionnaire. Sample size calculations were done for longitudinal analyses of changes in the primary outcome, PA patterns, yielding approximately 400 participants.^17^ Informed consent was obtained by all participants prior to participation.

### 2.4 Questionnaire

The online questionnaire was created in German and distributed with Research Electronic Data Capture (REDCap).^18, 19^ It included items gathering information about subject characteristics (including COVID-19 history and vaccination status), socio-economic data, subjective PA, barriers to PA, and objectively measured PA.

Subjective PA was assessed by the Global Physical Activity Questionnaire (GPAQ) consisting of 16 items about work, transport and leisure PA as well as sitting time.^20^ Weekly metabolic equivalent of task (MET)-minutes per week were calculated for total PA (TPA), vigorous PA (VPA), and moderate PA (MPA). Data processing and analyses of the GPAQ were done according to the guidelines of the World Health Organisation.^21^ Of note, previous findings showed that the use of absolute intensity cut-offs leads to the false classification of PA in populations other than those the cut-offs were derived from.^22^ Thus, age-adjusted MET-values referring to Garber et al.^23^ were used in this study (Table 1).

**Table 1.** Age-adjusted MET-values for moderate and vigorous physical activity.

| Age groups | METs |  |
| --- | --- | --- |
|  | Moderate | Vigorous |
| 18-39 | 7 | 9 |
| 40-64 | 6 | 7.5 |
| ≥65 | 4.5 | 5.75 |

Abbreviations: MET = metabolic equivalent of task (1 MET = 3.5 mL·kg<sup>-1</sup>·min<sup>-1</sup>).

For additional details regarding the type of sports, participants were asked to provide the most and second most frequent type of sports for both moderate and vigorous intensity. An adapted version of a questionnaire by Reichert et al.^24^ was utilized to gather information about perceived barriers to PA including three COVID-19-specific items (i.e. the anxiety of infection, lack of space, and restricted due to safety measures).

### 2.5 Data analysis

Statistics and figures were done in R version 4.0.3 (R Foundation for Statistical Computing, Vienna, Austria).^25^ Vaccination status was categorized as follows: unvaccinated, received first shot but not fully vaccinated, fully vaccinated, as well as recovered from COVID-19 within the last 6 months and unvaccinated. For moderate and vigorous PA, a considerable fraction of participants reported zero activity. Due to the clumping at zero, we used a two-step procedure^26-28^ to explore the association between COVID-19 vaccination status and PA patterns. During the first step, multiple logistic regressions were run with the respective PA pattern as a dichotomous dependent variable (engaged in PA: yes/no). During the second step, we used multiple linear regression models on the non-zero part where the log-transformed PA pattern served as the dependent variable and the dummy-coded categorical vaccination status served as the focal predictor. To minimize the bias of the estimate of the association between vaccination status and physical activity, we constructed a directed acyclic graph (DAG)^29^ which encodes our assumptions about the causal structure between the variables (see Supplementary File). Based on this DAG, all our models were adjusted for age, body mass index, sex (m/f), education level (see Table 1), living with others (yes/no), and area of residence (city center, suburbs, countryside). Age and body mass index were included with restricted cubic splines with four knots placed at appropriate percentiles of the data.^30^ This was done for PA patterns where we saw a considerable share of subjects reporting no PA. Residual diagnostics were used to verify that the model assumptions were satisfied. Pairwise between-group differences were further examined using pre-planned contrasts which were adjusted for multiple testing. Because we analyzed PA patterns on the log-scale, the estimated marginal means of the vaccination status groups represent geometric means. Hence, PA patterns between vaccination groups were compared using the ratio of their geometric means. Only for sedentary time we compared vaccination groups using differences between means. Sensitivity analyses were run for the main analyses additionally including the country of residence as a parameter in the models. Between-group differences in sporting behavior, as well as frequencies of perceived barriers to PA, were examined using Pearson’s Chi-square test. If any cells had expected frequencies below 5, Fisher’s exact test was used instead. For these tests, the false-discovery rate was controlled at the 5% level using the Benjamini-Hochberg method.^31^ Subsequently, post-hoc analyses were performed. All tests were two-sided and the statistical significance level was set to .05.

## 3. Results

### 3.1 Study cohort

Out of 2454 responses to the questionnaire, 1516 were complete and ended up being included in the analyses. Subject characteristics stratified by COVID-19 vaccination status are displayed in Table 2.

**Table 2.** Subject characteristics stratified by COVID-19 vaccination status.

| Variable | Total sample<br>(n = 1516) | Unvaccinated<br>(n = 157) | First short<br>(n = 48) | Fully<br>vaccinated (n<br>= 1269) | Recovered<br>(n = 42) |
| --- | --- | --- | --- | --- | --- |
| Female sex | 70.1 | 72.0 | 70.8 | 69.8 | 71.4 |
| Age, years <sup>\$</sup> | 41 (33) | 38 (30) | 32 (37) | 42 (33) | 38 (30) |
| BMI, kg·m<sup>-2</sup> | 24.0 ± 4.2 | 23.5 ± 3.9 | 22.6 ± 3.3 | 24.1 ± 4.3 | 24.3 ± 3.8 |
| **Country of residence** |  |  |  |  |  |
| Switzerland | 38.1 | 43.3 | 35.4 | 37.4 | 42.9 |
| Germany | 38.0 | 28.7 | 35.4 | 39.3 | 35.7 |
| Austria | 23.8 | 28.0 | 29.2 | 23.2 | 21.4 |
| **Level of education** |  |  |  |  |  |
| Vocational school | 16.4 | 21.2 | 16.7 | 15.7 | 11.9 |
| Secondary school | 11.8 | 14.6 | 12.5 | 11.1 | 21.4 |
| A-levels | 21.0 | 21.0 | 29.1 | 20.6 | 23.8 |
| University | 50.7 | 41.4 | 41.7 | 52.4 | 42.9 |
| **Civil status** |  |  |  |  |  |
| Single | 32.1 | 34.4 | 39.6 | 31.5 | 38.1 |
| Married | 39.5 | 40.8 | 33.3 | 39.3 | 45.2 |
| Divorced | 5.5 | 8.9 | 8.3 | 5.1 | 2.4 |
| Widowed | 2.7 | 1.3 | 2.1 | 2.9 | 2.4 |
| Relationship | 12.9 | 14 | 16.7 | 21 | 11.9 |
| Smoking, yes | 7.7 | 7.6 | 2.1 | 7.8 | 11.9 |
| Living w/<br>others, yes | 76.5 | 80.3 | 79.2 | 75.9 | 78.6 |

Data are presented as mean $\pm$ SD or relative frequencies unless denoted otherwise. The education levels ‘primary school’ and ‘vocational school’ were combined as the former made up only 0.5% in the full sample. Abbreviations: $\$$ = median (interquartile range); BMI = body-mass index.

### 3.2 Association between vaccination status and PA patterns

There was little evidence for between-group differences regarding the probability of being active regarding the respective PA patterns (Table 3).

**Table 3.** Two-part models for the association of physical activity patterns with vaccination status.

| Group comparisons | Logistic regression |  |  | Linear model |  |  |
| --- | --- | --- | --- | --- | --- | --- |
|  | OR (no PA) | 95% *CI* | *p*-value | Ratio | 95% *CI* | *p*-value |
| **TPA** |  |  |  |  |  | **0.011** |
| UV vs. 1V | - | - | - | 1.34 | 0.82 to 2.2 | 0.409 |
| UV vs. FV | - | - | - | 1.22 | 0.99 to 1.51 | 0.076 |
| UV vs. R | - | - | - | 1.05 | 0.69 to 1.58 | 0.991 |
| 1V vs. FV | - | - | - | 0.91 | 0.58 to 1.44 | 0.946 |
| 1V vs. R | - | - | - | 0.78 | 0.44 to 1.39 | 0.679 |
| FV vs. R | - | - | - | 0.86 | 0.60 to 1.24 | 0.703 |
| **VPA<sup>b</sup>** |  |  | 0.500 |  |  | **0.015** |
| UV vs. 1V | 0.56 | 0.21 to 1.46 | 0.395 | 1.60 | 0.99 to 2.58 | 0.057 |
| UV vs. FV | 0.89 | 0.52 to 1.50 | 0.932 | 1.09 | 0.86 to 1.39 | 0.763 |
| UV vs. R | 0.99 | 0.34 to 2.89 | 1.000 | 1.2 | 0.82 to 1.75 | 0.596 |
| 1V vs. FV | 1.59 | 0.68 to 3.72 | 0.488 | 0.68 | 0.45 to 1.05 | 0.104 |
| 1V vs. R | 1.77 | 0.50 to 6.28 | 0.640 | 0.75 | 0.45 to 1.26 | 0.479 |
| FV vs. R | 1.12 | 0.43 to 2.94 | 0.991 | 1.10 | 0.80 to 1.50 | 0.865 |
| **MPA<sup>c</sup>** |  |  | 0.089 |  |  | **0.001** |
| UV vs. 1V | 0.32 | 0.09 to 1.17 | 0.057 | 0.75 | 0.44 to 1.29 | 0.520 |
| UV vs. FV | 0.5 | 0.21 to 1.20 | 0.763 | 1.23 | 0.96 to 1.57 | 0.145 |
| UV vs. R | 0.42 | 0.10 to 1.70 | 0.596 | 0.74 | 0.46 to 1.17 | 0.320 |
| 1V vs. FV | 1.59 | 0.57 to 4.44 | 0.104 | 1.62 | 1.00 to 2.65 | 0.053 |
| 1V vs. R | 1.32 | 0.29 to 5.98 | 0.479 | 0.97 | 0.52 to 1.83 | 1.000 |
| FV vs. R | 0.83 | 0.26 to 2.64 | 0.865 | 0.60 | 0.40 to 0.90 | **0.009** |
| **Transport<sup>d</sup>** |  |  | 0.067 |  |  | 0.431 |
| UV vs. 1V | 2.97 | 0.90 to 9.79 | 0.086 | 1.26 | 0.86 to 1.85 | 0.383 |
| UV vs. FV | 1.07 | 0.67 to 1.71 | 0.981 | 1.15 | 0.91 to 1.46 | 0.404 |
| UV vs. R | 0.92 | 0.36 to 2.37 | 0.996 | 1.17 | 0.72 to 1.91 | 0.839 |
| 1V vs. FV | 0.36 | 0.12 to 1.11 | 0.088 | 0.91 | 0.66 to 1.26 | 0.880 |
| 1V vs. R | 0.31 | 0.08 to 1.24 | 0.131 | 0.93 | 0.54 to 1.58 | 0.982 |
| FV vs. R | 0.86 | 0.37 to 2.02 | 0.968 | 1.02 | 0.65 to 1.59 | 1.000 |

<sup>a</sup> n = 1478, <sup>b</sup> n = 1510, <sup>c</sup> n = 1260, <sup>d</sup> n = 1510. Abbreviations: FV = fully vaccinated; MPA = moderate physical activity; OR = odds ratio; R = recovered from coronavirus disease 2019 within the last 6 months; TPA = total physical activity; Transport = transport-related physical activity; UV = unvaccinated; 1V = first-shot; 95% *CI* = 95% confidence interval.

Overall, we found evidence for between-group differences in TPA (n = 1478, χ^2^(6) = 16.632, *p* = .011), VPA (n = 1109, χ^2^(6) = 15.819, *p* = .015), and MPA (n = 1339, χ^2^(6) = 21.764, *p* = .001) (see Table 3). However, there were no significant differences for transport-related PA (n = 1062, χ^2^(6) = 5.918, *p* = .432; see Table 3) and sedentary time (n = 1510, χ^2^(6) = 7.512, *p* = .276) between groups (Table 4).

**Table 4.** Linear model for sedentary time with between-group comparisons.

| **Group comparisons** | **Estimate** | **95% *CI*** | ***p*-value** |
| --- | --- | --- | --- |
|  |  |  | 0.276 |
| UV vs. 1V | 7.82 | -70.40 to 86.03 | .994 |
| UV vs. FV | -33.40 | -73.65 to 6.85 | .139 |
| UV vs. R | -19.74 | -110.09 to 70.60 | .940 |
| 1V vs. FV | -41.22 | -111.16 to 28.72 | .418 |
| 1V vs. R | -27.56 | -134.49 to 79.37 | .907 |
| FV vs. R | 13.66 | -69.61 to 96.94 | .97 |

Abbreviations: FV = fully vaccinated; R, recovered from coronavirus disease 2019 within the last 6 months; UV = unvaccinated; 1V = first-shot; 95% *CI* = 95% confidence interval.

### 3.3 Association between vaccination status and contribution of PA category to TPA

The relative contributions of work-, leisure time-, and transport-related PA to TPA were not significantly different between groups (Supplementary File).

### 3.4 Association between vaccination status and sporting behavior

The association of sporting behavior with vaccination status is presented in Figure 1. We found no evidence for between-group differences in any of the sports categories except for jogging as the most frequent intensive type of sports (*p* = .027). Individuals that had COVID-19 within the past six months were significantly different (*p* = .010).

**Fig 1.**
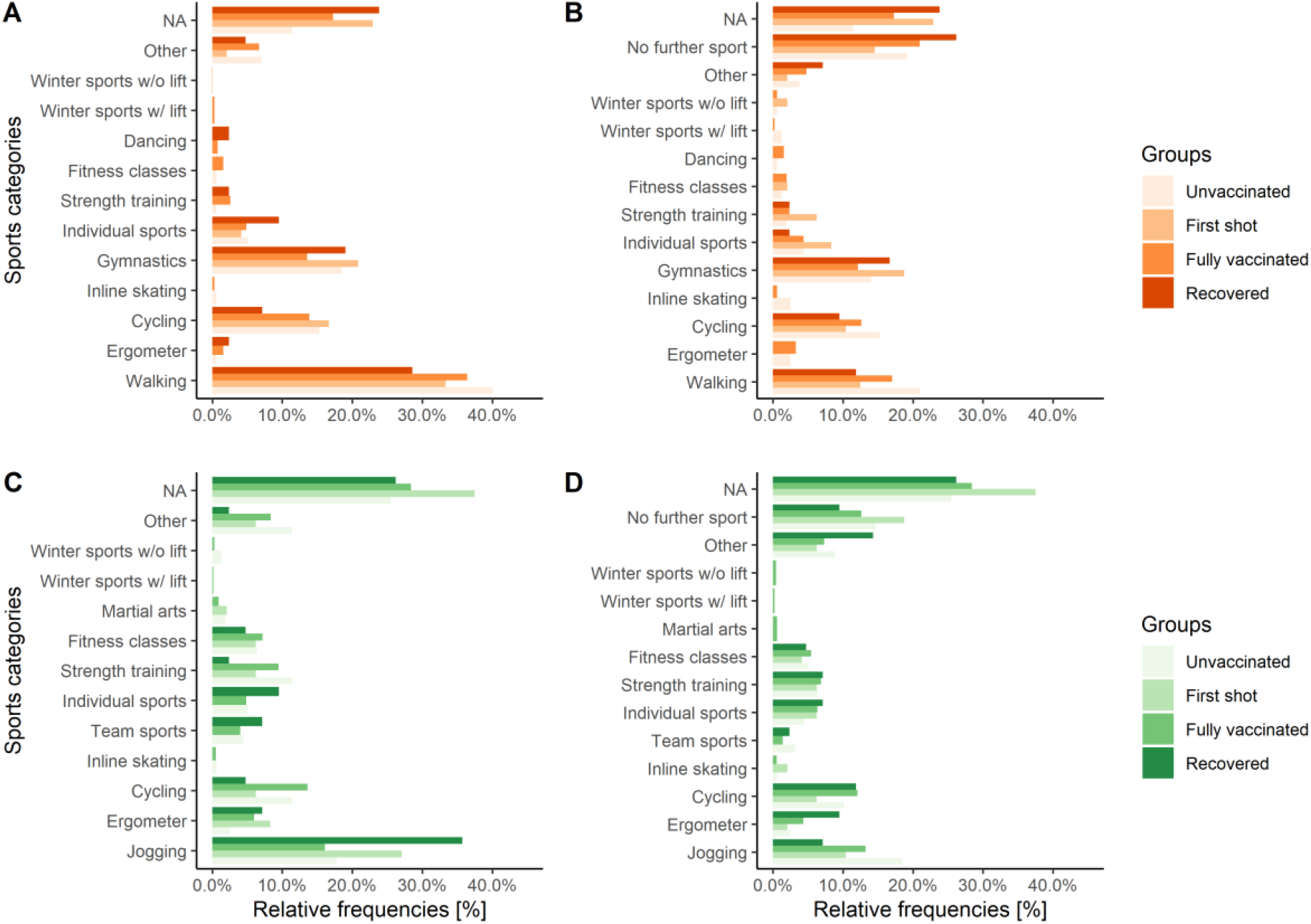
Frequencies of types of sports. A and B represent the most and second most frequent types of intensive sports, whereas C and D show the most- and second-most frequent types of moderate sports. No multiple choices per panel were possible. NA = no sports.

### 3.5 Association between vaccination status and barriers to PA

The results regarding barriers to PA are presented in Figure 2. Significant between-group differences were found for ‘current injury/disease’ (*p* < .001). Unvaccinated (*p* < .0001) and fully vaccinated individuals (*p* < .001) differed significantly. No evidence for differences was found for any of the COVID-19 specific factors.

**Fig 2.**
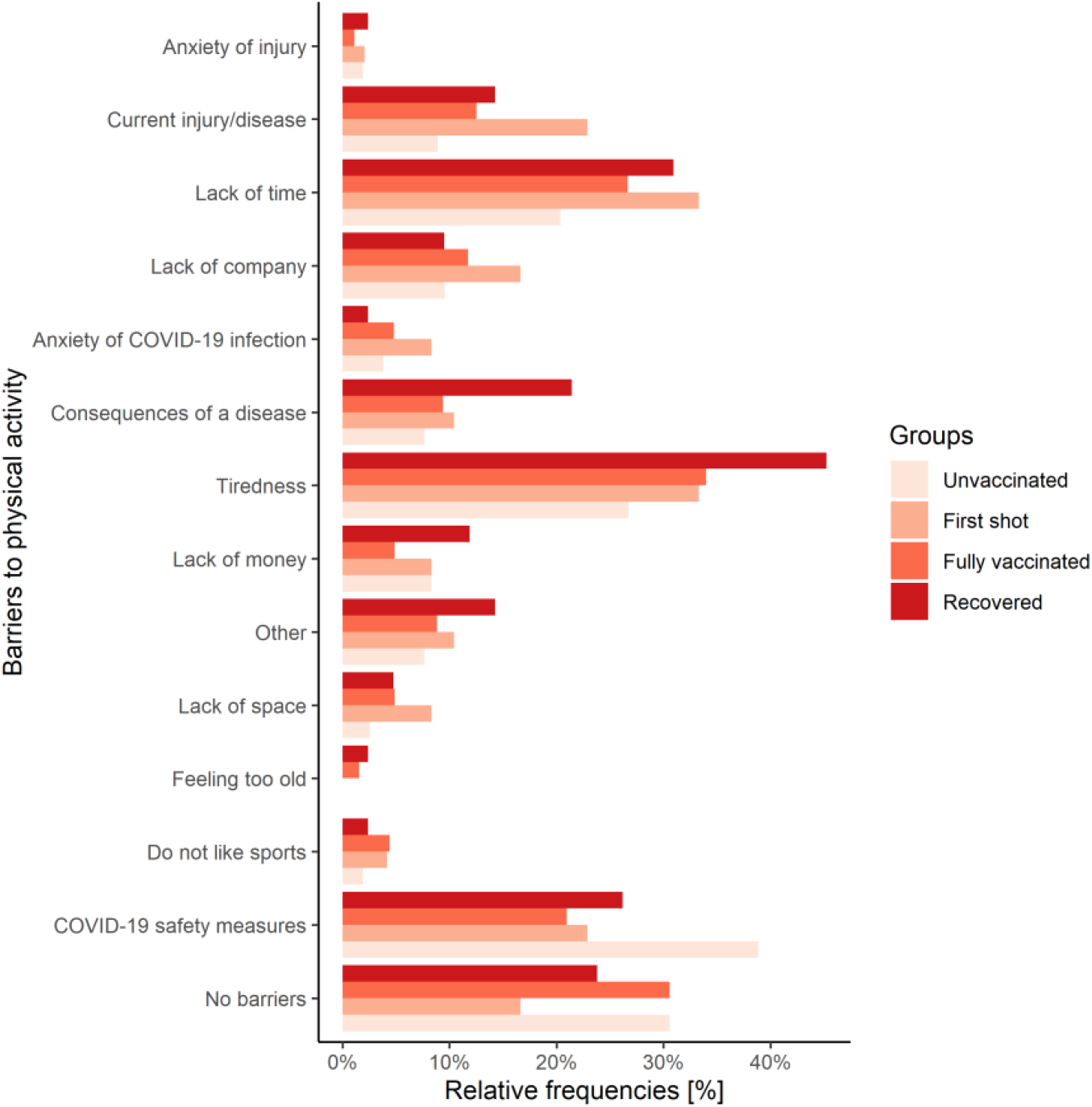
Frequencies of barriers to physical activity stratified by COVID-19 vaccination status. Multiple choices were possible. Abbreviations: COVID-19 = coronavirus disease 2019.

### 3.6 Sensitivity analyses

Results did not substantially differ when including the country of residence as an additional predictor into the main models.

## 4. Discussion

The major novel findings of this study were that firstly, COVID-19 vaccination status was associated with TPA, VPA, as well as MPA but not transport-related PA and sedentary time. Secondly, there was no evidence for a difference in the contribution of PA categories to TPA between vaccination status. Thirdly, we found no evidence that vaccination status was associated with sporting behavior except for jogging as a primary type of intense sport. Finally, between-group differences in barriers to PA were only apparent for ‘current injury/disease’ but not for COVID-19-specific factors.

### 4.1 Association between vaccination status and PA patterns

Overall, vaccination status is related to TPA, VPA and MPA. These findings are especially interesting considering that the survey was conducted in August 2021. Then, the occurrence of infections was low and safety measures were mostly mild, hence not substantially impacting individuals in their daily activities. In detail, our results indicated that unvaccinated individuals had higher levels of TPA and VPA but not MPA than individuals vaccinated once. Similar findings were apparent for TPA, VPA, but not MPA when comparing unvaccinated with fully vaccinated individuals. Yet, there was not sufficient evidence to verify these findings. Nonetheless, the discoveries are rather surprising and encourage further research in this area. Against our original assumption of vaccinated individuals being more engaged in PA due to being less restricted by safety measures in place and feeling more protected, we propose the following potential explanation. On one hand, vaccinated individuals might be more prudent to avoid infection and have different health awareness. On the other hand, unvaccinated individuals might feel more protected by a better immune system as a consequence of generally higher PA levels. This could be supported by recent research highlighting the beneficial effect of PA on the immune response^32, 33^ as well as the odds for hospitalization due to COVID-19.^8^ Also notable is that individuals with COVID-19 in the past 6 months engaged in more TPA than both partly and fully vaccinated individuals. Again, there was not sufficient evidence to confirm this association. However, for MPA significantly higher levels could be seen than in fully vaccinated individuals. Thinking of highly prevalent symptoms of post-COVID-19 syndrome^34, 35^ such as exercise intolerance and reduced resilience or simply the deconditioning experienced after a period of bed rest, these individuals might be dedicated to getting back in shape or counteracting residual symptoms. The abovementioned findings, taken together, highlight the role of vaccination status as an explanatory variable for changes in PA patterns in the current situation. Moreover, if these differences in PA should persist over time, health benefits/detriments for some individuals depending on their vaccination status might be observable.^36^ Finally, there was only little evidence for the association of transport-related PA and sedentary time with vaccination status. This was expected since for these PA patterns safety measures equally apply to all groups.

### 4.2 Association between vaccination status and contribution of PA category to TPA

To better understand the origin of alteration in TPA, it is useful to analyze the contribution of the respective PA categories to TPA. The relative contributions of PA categories, i.e. work-, leisure time-, and transport-related PA, were not different between groups. This indicates that the between-group differences seen in TPA might not be due to a single PA category. It thus seems relevant to examine all categories of PA when searching for the main contributor to between-group differences in TPA.

### 4.3 Association between vaccination status and sporting behavior

Interestingly, despite the role of vaccination status in TPA, VPA, and MPA, sporting behavior seems not to be associated with vaccination status except jogging as the most frequent type of intensive sport. Individuals with COVID-19 in the past 6 months reported to engage more frequently in jogging than the other groups. This aligns with the finding of those with COVID-19 in the past performing more MPA than fully vaccinated individuals. Jogging is among the most popular types of sports in Europe, is easily accessible, requires minimal equipment, and does not necessitate contact with others.^37^ For those reasons it might be the go-to activity for getting back in shape post-infection. On the contrary, since the other groups did not differ, this could mean that their sporting behavior is more diverse. Yet, detailed conclusions are difficult to draw from the present data. Detailed data on the time spent in the respective types of sports would be valuable.

### 4.4 Association between vaccination status and barriers to PA

Barriers to PA describe what prevents individuals from being physically active. Interestingly, there was only little evidence for between-group differences regarding the COVID-19-specific barriers. Yet, unvaccinated individuals tended to report ‘COVID-19 safety measures’ more often, whereas among individuals with COVID-19 in the past, ‘tiredness’ and ‘consequences of a disease’ tended to be more frequently reported. The latter two barriers are in line with data on post-COVID-19 syndrome.^34, 35^ However, the lack of evidence may be explained by the circumstances the survey was conducted in – low occurrence of infections, mild safety measures, and for instance in Switzerland no vaccination certificate mandate at the time.^38^

### 4.5 Sensitivity analyses

We found that the association between PA and vaccination status was not altered by the country of residence. This was expected. Safety measures^15^, the fraction of fully-vaccinated individuals, as well as the number of new infections was similar in all three countries.^16^ However, the country of residence may be relevant in survey round 2 that is being conducted at the moment (December 2021). At the time, there was a countrywide lockdown in Austria whereas this was not the case for Germany and Switzerland.^15^ The present transnational study will be of great value in such situations.

### 4.6 Strengths and limitations

The main strength of this study is its transnational prospective study design. It allows observing the impact of the situation at the time (e.g. differences in severity of implemented safety measures and number of new infections) between the respective countries on the aforementioned associations. Together with the results of survey round 2 and 3, the influence of different time points (i.e. August & December 2021, August 2022) on these associations can likewise be observed. Another strength is the application of innovative statistical methods (i.e. two-step procedure^26-28^) in the area of sports medicine.

Several potential limitations should be discussed. Due to the design of the study, the group sizes were not balanced. This may be a reason for not reaching statistical significance in some of the post-hoc analyses. Moreover, the study cohort may likely not be representative of the German-speaking general population, especially because of the high percentage of female participants and individuals with a university degree. Nonetheless, the results are adaptable to a population of well-educated individuals.

## 5. Conclusion

To the best of our knowledge, this is the first study to investigate the association of vaccination status with PA. We showed that COVID-19 vaccination status is associated with PA levels in German-speaking countries and should thus be assessed in future research in this pandemic situation. There was little evidence for a clear contribution of a single PA category (work, leisure time, or transport) to changes in TPA. This highlights the need for assessing PA in its wide spectrum. Sporting behavior may not be associated with vaccination status. Only jogging was more frequently reported in individuals recovering from COVID-19 as it may be an easily accessible activity to get back in shape. Finally, barriers to PA were not significantly different between groups which may be explained by the low number of COVID-19 cases and mild safety measures at the time of the survey.

We recommend that COVID-19 vaccination status should be considered as a potential moderator for PA patterns in both future research and the development of PA guidelines in pandemic times. It remains to be seen whether these associations change in winter with likely stricter safety measures in place and rising numbers of infections. Survey round 2 of the COR-PHYS-Q study will investigate this.

## Data Availability

Data will be made available by the corresponding author upon reasonable request.

## Acknowledgements

The authors thank everyone that contributed to distributing the link to the questionnaire. Also, we are grateful for all individuals that participated in the COR-PHYS-Q study. No funding was received. Participants and the public were not involved in the planning or conduct of this study.

## Author’s contributions

FS conceptualized the study, defined the methods, collected the data, was responsible for data curation, analyzed and interpreted the data, and wrote the original draft of the manuscript. HTB contributed to collecting the data, writing, and revised the manuscript. MF contributed to collecting the data and revised the manuscript. UT contributed to define the methods and revised the manuscript. AST conceptualized the study, defined the methods, collected the data, and revised the manuscript. All authors have read and approved the final version and agree with the order of presentation of the authors.

## References

1. Stockwell S, Trott M, Tully M, et al. Changes in physical activity and sedentary behaviours from before to during the COVID-19 pandemic lockdown: A systematic review. BMJ Open Sport Exerc Med 2021;7:e000960.

2. Lee IM, Shiroma EJ, Lobelo F, Puska P, Blair SN, Katzmarzyk PT. Effect of physical inactivity on major non-communicable diseases worldwide: An analysis of burden of disease and life expectancy. Lancet 2012;380:219–29.

3. Bull FC, Al-Ansari SS, Biddle S, et al. World Health Organization 2020 guidelines on physical activity and sedentary behaviour. Br J Sports Med 2020;54:1451.

4. Ding D, Lawson KD, Kolbe-Alexander TL, et al. The economic burden of physical inactivity: A global analysis of major non-communicable diseases. Lancet 2016;388:1311–24.

5. World Health Organization. Stay physically active during self-quarantine; 2021. Available from: https://www.euro.who.int/en/health-topics/health-emergencies/coronavirus-COVID-19/publications-and-technical-guidance/noncommunicable-diseases/stay-physically-active-during-self-quarantine. [Accessed 09.12.2021].

6. National Center for Chronic Disease Prevention and Health Promotion. How to be physically active while social distancing; 2021. Available from: https://www.cdc.gov/physicalactivity/how-to-be-physically-active-while-social-distancing.html. [Accessed 09.12.2021].

7. Laddu DR, Lavie CJ, Phillips SA, Arena R. Physical activity for immunity protection: Inoculating populations with healthy living medicine in preparation for the next pandemic. Prog Cardiovasc Dis 2021;64:102–4.

8. Sallis R, Young DR, Tartof SY, et al. Physical inactivity is associated with a higher risk for severe COVID-19 outcomes: A study in 48 440 adult patients. Br J Sports Med 2021;55:1099–105.

9. Campbell JP, Turner JE. Debunking the myth of exercise-induced immune suppression: Redefining the impact of exercise on immunological health across the lifespan. Front Immunol 2018;9. doi: 10.3389/fimmu.2018.00648

10. Polack FP, Thomas SJ, Kitchin N, et al. Safety and efficacy of the BNT162b2 mRNA COVID-19 vaccine. New Engl J Med 2020;383:2603–15.

11. Voysey M, Clemens SAC, Madhi SA, et al. Safety and efficacy of the ChAdOx1 nCoV-19 vaccine (AZD1222) against SARS-CoV-2: An interim analysis of four randomised controlled trials in brazil, south africa, and the uk. Lancet 2021;397:99–111.

12. Mahase E. COVID-19: Moderna applies for US and EU approval as vaccine trial reports 94.1% efficacy. BMJ 2020;371:m4709.

13. Mathieu E, Ritchie H, Ortiz-Ospina E, et al. A global database of COVID-19 vaccinations. Nat Hum Behav 2021;5:947–53.

14. Ghram A, Moalla W, Lavie CJ. Vaccine and physical activity in the era of COVID-19 pandemic. Prog Cardiovasc Dis 2021;67:33–4.

15. Hale T, Sam W., Anna P., Toby P. K.B., Oxford COVID-19 government response tracker. In: Government BSo, ed.: Blavatnik School of Government; 2020.

16. Dong E, Du H, Gardner L. An interactive web-based dashboard to track COVID-19 in real time. Lancet Infect Dis 2020;20:533–4.

17. Cheval B, Sivaramakrishnan H, Maltagliati S, et al. Relationships between changes in self-reported physical activity, sedentary behaviour and health during the coronavirus (COVID-19) pandemic in France and Switzerland. J Sports Sci 2020;39:699–704.

18. Harris PA, Taylor R, Minor BL, et al. The redcap consortium: Building an international community of software platform partners. J Biomed Inform 2019;95:103208.

19. Harris PA, Taylor R, Thielke R, Payne J, Gonzalez N, Conde JG. Research electronic data capture (REDCap) – a metadata-driven methodology and workflow process for providing translational research informatics support. J Biomed Inform 2009;42:377–81.

20. Wanner M, Hartmann C, Pestoni G, Martin BW, Siegrist M, Martin-Diener E. Validation of the global physical activity questionnaire for self-administration in a european context. BMJ Open Sport Exerc Med 2017;3:e000206.

21. World Health Organization. Global Physical Activity Questionnaire (GPAQ): Analysis guide; Available from: https://www.who.int/ncds/surveillance/steps/resources/GPAQ_Analysis_Guide.pdf. [Accessed 01.10.2021].

22. Schwendinger F, Wagner J, Infanger D, Schmidt-Trucksäss A, Knaier R. Methodological aspects for accelerometer-based assessment of physical activity in heart failure and health. BMC Med Res Methodol 2021;21:251.

23. Garber CE, Blissmer B, Deschenes MR, et al. American college of sports medicine position stand. Quantity and quality of exercise for developing and maintaining cardiorespiratory, musculoskeletal, and neuromotor fitness in apparently healthy adults: Guidance for prescribing exercise. Med Sci Sports Exerc 2011;43:1334–59.

24. Reichert FF, Barros AJD, Domingues MR, Hallal PC. The role of perceived personal barriers to engagement in leisure-time physical activity. Am J Public Health 2007;97:515–9.

25. R Core Team. R: A language and environment for statistical computing; 2021. Available from: https://www.R-project.org/.

26. Min Y, Agresti A. Modeling nonnegative data with clumping at zero: A survey. J of Iran Stat Soc 2002;1:7–33.

27. Boulton AJ, Williford A. Analyzing skewed continuous outcomes with many zeros: A tutorial for social work and youth prevention science researchers. J Soc Soc Work and Res 2018;9:721–40.

28. Neelon B, O’Malley AJ, Smith VA. Modeling zero-modified count and semicontinuous data in health services research part 2: Case studies. Stat Med 2016;35:5094–112.

29. Shrier I, Platt RW. Reducing bias through directed acyclic graphs. BMC Med res Methodol 2008;8:70.

30. Harrell FE. Regression modeling strategies: With applications to linear models, logistic and ordinal regression, and survival analysis. 2nd ed.: Springer; 2016.

31. Benjamini Y, Hochberg Y. Controlling the false discovery rate: A practical and powerful approach to multiple testing. J R Stat Soc Series B 1995;57:289–300.

32. Burtscher J, Burtscher M, Millet GP. The central role of mitochondrial fitness on antiviral defenses: An advocacy for physical activity during the COVID-19 pandemic. Redox Biol 2021;43:101976.

33. Burtscher J, Millet GP, Burtscher M. Low cardiorespiratory and mitochondrial fitness as risk factors in viral infections: Implications for COVID-19. Br J Sports Med 2020;55:413–15.

34. Nehme M, Braillard O, Chappuis F, Courvoisier DS, Guessous I. Prevalence of symptoms more than seven months after diagnosis of symptomatic COVID-19 in an outpatient setting. Ann Intern Med 2021;174:1252–60.

35. Gebhard CE, Sütsch C, Bengs S, et al. Sex- and gender-specific risk factors of post-COVID-19 syndrome: A population-based cohort study in Switzerland. medRxiv 2021:2021.06.30.21259757. doi: 10.1101/2021.06.30.21259757

36. Lavie CJ, Ozemek C, Carbone S, Katzmarzyk PT, Blair SN. Sedentary behavior, exercise, and cardiovascular health. Circ Res 2019;124:799–815.

37. Hulteen RM, Smith JJ, Morgan PJ, et al. Global participation in sport and leisure-time physical activities: A systematic review and meta-analysis. Prev Med 2017;95:14–25.

38. Federal Office of Public Health (FOPH). COVID-19 Switzerland - information on the current situation, as of 10 September 2021; 2021. Available from: https://www.covid19.admin.ch/en/epidemiologic/hosp. [Accessed 09.09.2021].

