## Supplementary File for "COVID-19 vaccination status is associated with physical activity in German-speaking countries: the COR-PHYS-Q cohort study"

Table 1
Logistic regression results of relative contributions of physical activity categories to total physical activity (n = 1478).

| Group comparisons | OR | 95% *CI* | *p*-value |
|---|---|---|---|
| **Work** | | | 0.612 |
| UV vs. 1V | 1.00 | 0.43 to 2.32 | 1.000 |
| UV vs. FV | 1.12 | 0.74 to 1.69 | 0.895 |
| UV vs. R | 0.68 | 0.33 to 1.42 | 0.525 |
| 1V vs. FV | 1.12 | 0.52 to 2.43 | 0.980 |
| 1V vs. R | 0.68 | 0.26 to 1.83 | 0.747 |
| FV vs. R | 0.61 | 0.32 to 1.16 | 0.191 |
| **Leisure time** | | | 0.572 |
| UV vs. 1V | 1.45 | 0.78 to 2.70 | 0.403 |
| UV vs. FV | 0.96 | 0.70 to 1.32 | 0.986 |
| UV vs. R | 1.13 | 0.62 to 2.08 | 0.951 |
| 1V vs. FV | 0.66 | 0.38 to 1.16 | 0.220 |
| 1V vs. R | 0.78 | 0.36 to 1.67 | 0.830 |
| FV vs. R | 1.18 | 0.69 to 2.02 | 0.855 |
| **Transport** | | | 0.434 |
| UV vs. 1V | 0.57 | 0.28 to 1.18 | 0.190 |
| UV vs. FV | 0.95 | 0.67 to 1.34 | 0.976 |
| UV vs. R | 1.35 | 0.65 to 2.80 | 0.703 |
| 1V vs. FV | 1.65 | 0.86 to 3.19 | 0.198 |
| 1V vs. R | 2.36 | 0.94 to 5.91 | 0.075 |
| FV vs. R | 1.43 | 0.74 to 2.76 | 0.495 |

Abbreviations: FV = fully vaccinated; leisure time = leisure-time related physical activity; OR = odds ratio; R = recovered from coronavirus disease 2019 within the last 6 months; Transport = transport-related physical activity; UV = unvaccinated; Work = work-related physical activity; 1V = first-shot; 95% *CI* = 95% confidence interval.

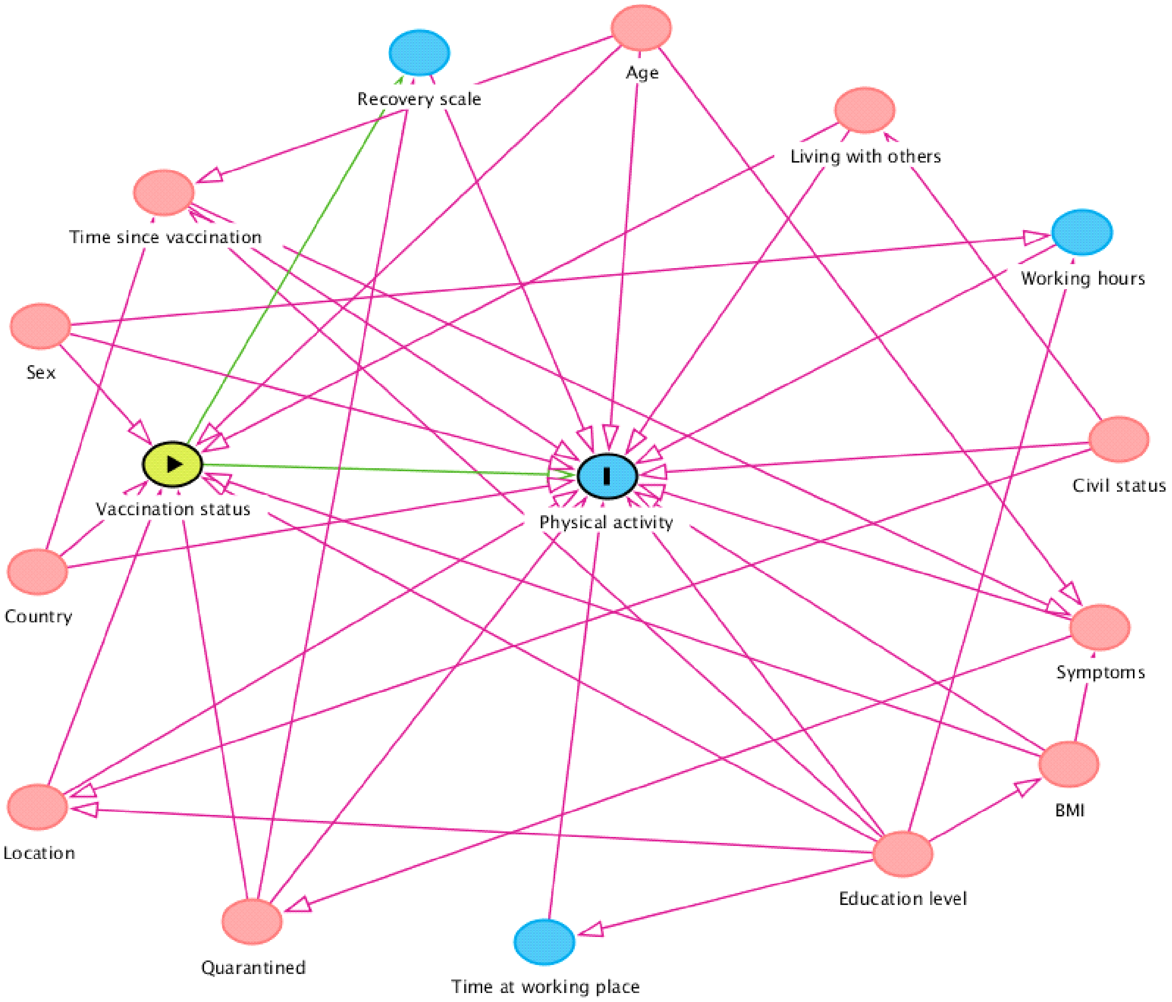


Fig 1. Directed acyclic graph (DAG). Abbreviations: BMI = body-mass index.
